# Detection of Neuron-Derived cfDNA in Blood Plasma: A New Diagnostic Approach for Neurodegenerative Conditions

**DOI:** 10.1101/2023.08.08.23293455

**Authors:** Chad A. Pollard, Kenneth Aston, Benjamin R. Emery, Jonathon Hill, Timothy Jenkins

## Abstract

Neurodegenerative diseases, such as Alzheimer’s disease (AD), pose significant challenges in early diagnosis, leading to irreversible brain damage and cognitive decline. In this study, we present a novel diagnostic approach that utilizes whole molecule analysis of neuron-derived cell-free DNA (cfDNA) as a biomarker for early detection of neurodegenerative diseases. By analyzing Differential Methylation Regions (DMRs) between purified cortical neurons and blood plasma samples, we identified robust biomarkers that accurately distinguish between neuronal and non-neuronal cfDNA. The use of cfDNA offers the advantage of convenient and minimally invasive sample collection compared to traditional cerebrospinal fluid or tissue biopsies, making this approach more accessible and patient friendly. Targeted sequencing at the identified DMR locus demonstrated that a conservative cutoff of 5% of neuron-derived cfDNA in blood plasma accurately identifies 100% of patients diagnosed with AD, showing promising potential for early disease detection. Additionally, this method effectively differentiated between patients with mild cognitive impairment (MCI) who later progressed to AD and those who did not, highlighting its prognostic capabilities. Importantly, the differentiation between patients with neurodegenerative diseases and healthy controls demonstrated the specificity of our approach. Furthermore, this cfDNA-based diagnostic strategy outperforms recently developed protein-based assays, which often lack accuracy and convenience. While our current approach focused on a limited set of loci, future research should explore the development of a more comprehensive model incorporating multiple loci to increase diagnostic accuracy further. Although certain limitations, such as technical variance associated with PCR amplification and bisulfite conversion, need to be addressed, this study emphasizes the potential of cfDNA analysis as a valuable tool for pre-symptomatic detection and monitoring of neurodegenerative diseases. With further development and validation, this innovative diagnostic strategy has the potential to significantly impact the field of neurodegenerative disease research and patient care, offering a promising avenue for early intervention and personalized therapeutic approaches.

## Introduction

Alzheimer’s disease, a devastating neurodegenerative disorder, poses formidable challenges in both diagnosis and treatment. Its pre-symptomatic detection remains elusive, and the current diagnostic approach primarily relies on documenting mental decline, leading to late-stage diagnosis when severe brain damage has already taken place. Compounding the issue, the disease’s gradual progression involves years of elevated neuronal cell death before visible symptoms emerge, with approximately 60 percent of neurons already lost by the time symptoms become apparent (1). Consequently, therapeutic interventions aiming to prevent the disease are rendered ineffective once symptoms manifest. Existing treatments offer modest relief, providing patients with only a few additional years of quality life before cognitive function significantly declines. However, early pre-symptomatic diagnosis has the potential to revolutionize disease management, allowing interventions that could halt the disease’s progression altogether, significantly improving patient outcomes.

At present, there are six drugs approved for treating mild to severe AD (2). Additionally, two disease-modifying immunotherapies have just recently received FDA accelerated approval for mild cognitive impairment or mild Alzheimer’s treatment (3), highlighting the need for better pre-symptomatic diagnosis with another 462 drugs currently under evaluation in clinical trials (2).

Recognizing the growing importance of pre-symptomatic diagnosis, considerable efforts have been directed towards developing biomarker-based approaches. Notable advancements in Alzheimer’s research have recently focused on the identification of A-beta 42 and A-beta 40 biomarkers in the blood (4). These biomarkers facilitate the detection of key pathologies, such as the abnormal buildup of amyloid and tau proteins, which are characteristic of AD (5). However, protein-based assays present challenges in terms of their complexity and requirement for precise conditions during sample handling to prevent protein denaturation (6). Further, this biomarker is most valuable only for the indication of AD specifically and not useful for other neurodegenerative conditions that could be addressed by an expansion of the approach assessed herein.

In this study, we propose a novel approach that circumvents the limitations of protein-based assays. It is a well-known fact that neuron-derived cfDNA circulates through the blood stream following neuron cell death (7). Our method leverages the power of DNA methylation analysis combined with whole molecule analysis via nanopore sequencing to identify and analyze this neuron-derived cfDNA. DNA methylation has emerged as a remarkable identifier of cell type (8) and its inherent stability allows for the potential to reduce costs and simplify sample processing (9). Moreover, by harnessing advanced sequencing technologies, we can achieve high accuracy in identifying rare DNA fragments such as dying neuronal DNA (10). Our approach focuses on identifying DNA methylation signatures and utilizing whole-molecule analysis to detect the presence of neuron-derived DNA molecules in the blood. The identification and monitoring of neuron-derived DNA serves as reliable predictors of neurodegenerative diseases, offering potential breakthroughs in the diagnosis and early intervention of not only AD, but all forms of neurodegenerative disease.

## Material and Methods

### Sample Acquisition

A total of 50 blood plasma samples were acquired from PrecisionMed, a company specializing in biospecimen procurement. The samples were carefully selected to include 25 older control individuals, 13 patients diagnosed with AD, 6 patients with mild cognitive impairment (MCI) who later progressed to confirmed AD, and 6 MCI patients who did not progress to AD. Age, sex, and ethnicity were matched across the sample groups to ensure comparability. Additionally, 10 blood plasma samples were obtained from Innovative Research as a young donor group, with an average age of 34. Furthermore, 4 technical replicates of purified cortical neurons were procured from Creative Bioarray and used as controls.

### Illumina Array Data Analysis

Publicly available Illumina Array EPIC data was retrieved from Gene Expression Omnibus (GEO) databases under the accession numbers GSE108462 (blood plasma samples) and GSE66351 (purified cortical neurons). Only control samples were utilized from both datasets. The raw IDAT methylation array data obtained from GEO datasets was preprocessed using the minfi R package. To obtain beta values for each cytosine-guanine dinucleotide (CpG), the data was SWAN normalized. Density plots of the beta value distribution were examined to ensure a bimodal nature, with prominent peaks between 0.0-0.2 and 0.8-1.0 and a flat valley between 0.2-0.8. Any samples deviating from this distribution were excluded, and the remaining samples were renormalized using the aforementioned procedures. Differentially Methylated Regions (DMRs) between neurons and blood plasma were identified using the University of Utah’s sliding window analysis tool (USeq), narrowing down the target sites for further analysis.

### Primer Design

Primers were designed using the online tool methprimer, specifically targeting bisulfite-converted DNA sequences. Due to the identical nature of CpG methylation on both the forward and reverse strands, primers were designed to amplify only the forward strands of the targeted loci. A 500-base pair window surrounding the targeted CpG was considered for primer design. To verify the effectiveness of the designed primers, electrophoretic gels were employed, and out of the 25 tested primers, 7 exhibited proper amplification, affirming their suitability for subsequent analysis.

### Sample Processing

cfDNA was extracted from each blood plasma sample using the Qiagen QIAamp MinElute ccfDNA Mini Kit (Catalog Number: 55204) following the provided protocol without modifications. DNA from purified cortical neurons was extracted using the Qiagen DNeasy Blood & Tissue Kit (Catalog Number: 69506) following the standard protocol. The samples were divided into two aliquots, enabling technical replication, resulting in a total of 120 blood samples and 8 purified cortical neuron samples. Following cfDNA extraction, bisulfite conversion was performed using the Zymo Research EZ DNA Methylation Kit (Catalog Number D5002), adhering to the recommended protocol. Subsequently, DNA was PCR amplified at two loci using the New England Biolabs 2X ZymoTaq Premix (Catalog Number: E2003-1). For each sample, the PCR reaction mixture consisted of 25 μl of 2X ZymoTaq Premix, 3 μl of the forward primer, 3 μl of the reverse primer, 17 μl of ultra-pure water, and 2 μl of bisulfite-converted DNA. The thermal cycling conditions were as follows: 1 cycle [95°C for 10 min], 38 cycles [95°C for 30 sec, 55°C for 30 sec, 72°C for 30 sec], and 1 cycle [72°C for 7 min]. The amplified loci targeted were chr3_42190778_42191048 and chr19_3507867_3507868. The forward and reverse primers for chr3_42190778_42191048 were TTTTATTGTTTTGGTTTTAGATTGT and AAATAAACTTCACAACACCATCAAC, respectively. The forward and reverse primers for chr19_3507867_3507868 were GGTATTATTTAGGTTTGGTTTT and TACCTTTAAATAAATATCTACTCCCTTAAC, respectively. Following amplification, the DNA library was prepared using the Nanopore SQK-NBD114.96 kit for short DNA fragments. One modification was made to the protocol by using a 1.0X concentration of AmpureXP beads instead of the recommended 0.4X to better target highly fragmented reads. The prepared libraries were then sequenced for 72 hours on the MinION MK1C Nanopore Sequencer, following the flow cell loading protocol provided with the SQK-NBD114.96 kit.

### Nanopore Data Analysis

All samples were sequenced using the MinION MK1C instrument with version 14 flow cells. Prior to sequencing, pore viability was checked for all flow cells to ensure optimal performance. Reads with a quality score of less than 9 were marked as failed and reads shorter than 200 bp were also excluded from analysis. Fastq files from each sample were analyzed using the microseq package in R. To focus on completely amplified molecules and avoid fragmented reads, only reads representing fully amplified molecules were utilized for subsequent analysis. Bisulfite conversion errors were checked by manually examining the reads for any C’s that were not in a CG context. Any reads with unconverted C’s outside a CG context were removed from the analysis. Whole-molecule analysis was then performed on each read for each sample by calculating the mean methylation across the molecule. Based on the methylation levels, the reads were categorized as blood-derived DNA (whole-molecule methylation > 0.75), neuron-derived DNA (whole-molecule methylation = 0), or unknown reads (whole-molecule methylation between 0 and 0.75). The ratio of neuron-derived DNA was calculated for each sample by dividing the number of neuron-derived reads by the total number of reads in the sample.

## Results

### Initial DMR Discovery via Array Data

A comprehensive sliding window analysis comparing purified cortical neurons and blood plasma samples revealed a total of 37,455 Differentially Methylated Regions (DMRs) with a minimum p-value of .00001. These DMRs represent potential markers for distinguishing between neuron-derived cfDNA and cfDNA in blood plasma. To visualize the distribution of these sites and highlight their discriminatory potential, a heat map was generated, vividly depicting the significant contrast between blood plasma and purified neurons (Figure 1).

**Figure 1:**
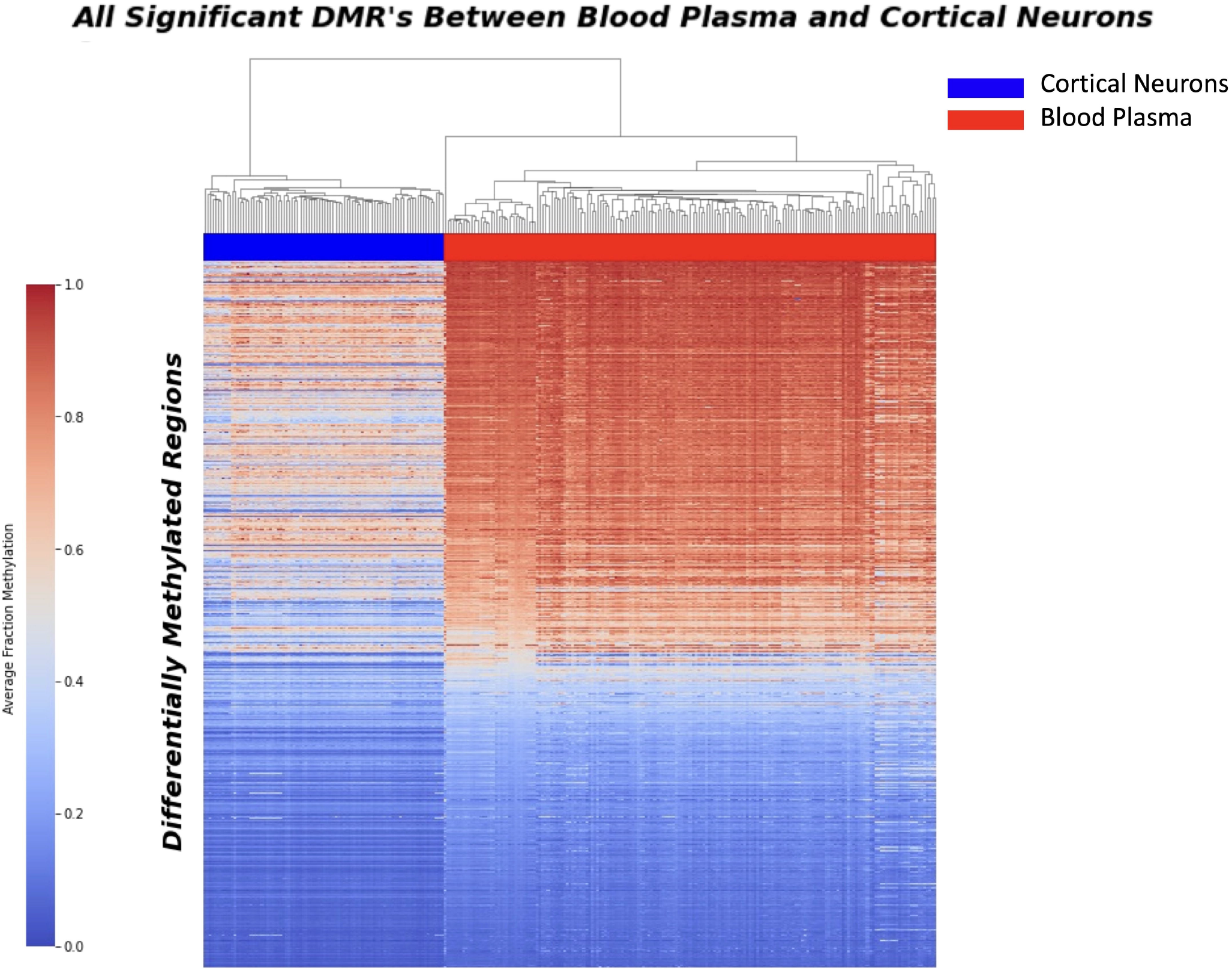
Heatmap displaying the distribution of 37,455 Differential Methylation Regions (DMRs) between purified neurons and blood plasma samples. The heatmap illustrates the methylation patterns at these DMRs, with red indicating higher methylation levels and blue representing lower methylation levels. Each row corresponds to a specific DMR, while each column represents a sample. The distinct clustering of samples based on their methylation patterns highlights the significant phenotypic difference between blood plasma and purified neurons, underscoring the potential of these DMRs as potential biomarkers for neurodegenerative disease detection.

To further focus our investigation on the most promising candidate sites, we conducted additional refinement on the identified DMRs. This refinement process led to the identification of 957 sites displaying an absolute mean difference of 0.6 or higher in methylation beta values. These selected sites showed substantial differences in methylation patterns between neurons and blood plasma, indicating their potential as robust markers for neuronal-derived cfDNA. The graphical representation of these differentially methylated sites is depicted in Figure 2.

**Figure 2:**
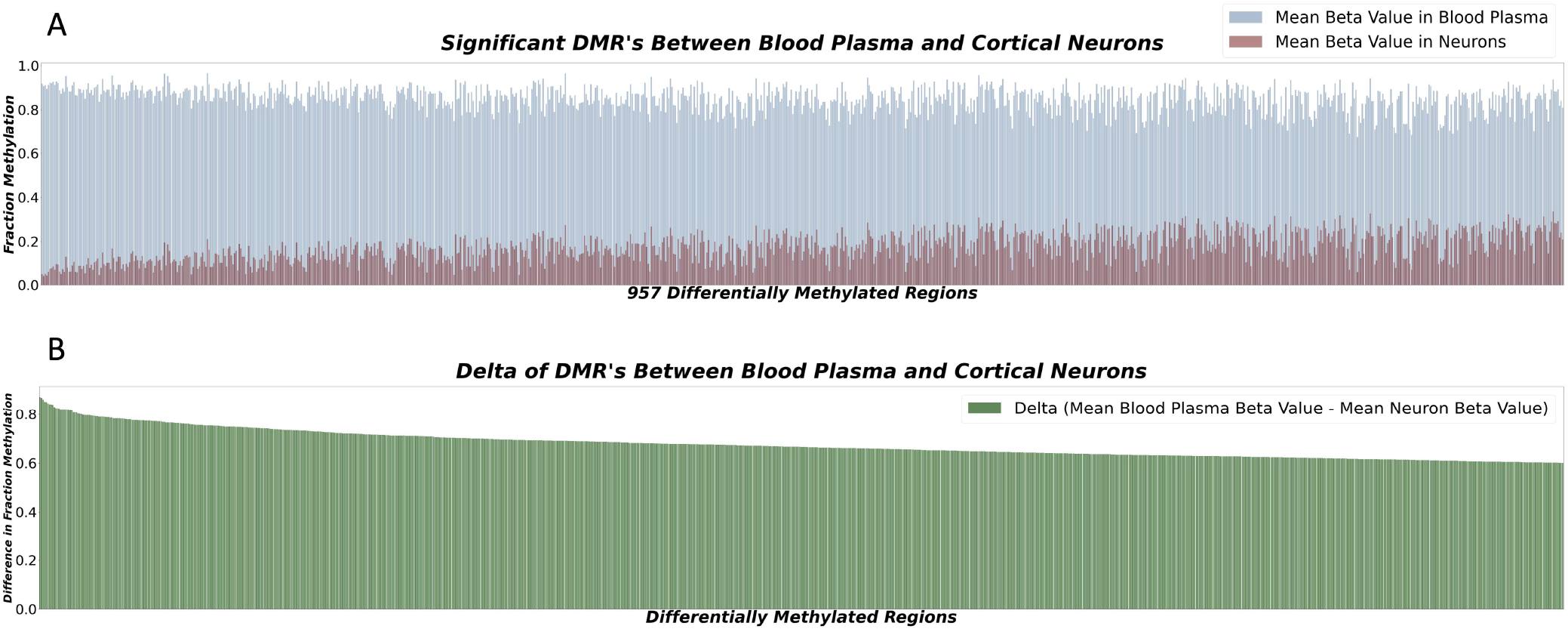
(A) Bar plot displaying 957 sites with a fraction methylation difference of ≥ 0.6 between cortical neurons and blood plasma. These sites show significant potential as biomarkers for neurodegenerative disease detection. (B) Delta representation of methylation differences between cortical neurons and blood plasma at selected loci. Positive values indicate higher methylation in neurons, while negative values signify lower methylation. These highly differentially methylated loci hold promise as accurate biomarkers for neuron-derived cfDNA, enhancing neurodegenerative disorder diagnostics.

### Whole-molecule Selection

To address the limited CpG representation on the Illumina EPIC array, we employed sequencing to explore the methylation status of the surrounding CpGs in our identified DMRs. After designing and optimizing primers for the top 25 candidate DMRs, seven primers were successfully amplified, confirming their suitability for subsequent sequencing analysis. Sequencing at the target loci was performed between 4 purified cortical neuron samples, and 4 pooled blood plasma samples.

Among the sequenced loci, all except Figure 3A demonstrated a significant separation (p-value ≤ 0.05) in methylation patterns across the whole DNA molecule between cortical neurons and blood plasma (Figure 3). Notably, the locus chr3:42190679,42191148 exhibited the strongest separation (p-value = 0.000042) across all CpGs of the entire DNA molecule. Specifically, purified neuron DNA reads displayed complete unmethylation at all CpG sites, while blood plasma DNA exhibited nearly full methylation at the same CpG sites (Figure 3B). This striking differential methylation pattern at chr3:42190679,42191148 underscored its potential as a promising biomarker for distinguishing neuronal-derived cell-free DNA (cfDNA) from cfDNA in blood plasma. Consequently, this locus was selected for comprehensive whole-molecule analysis in our acquired dataset, enabling its application in the diagnosis and monitoring of neurodegenerative diseases.

**Figure 3:**
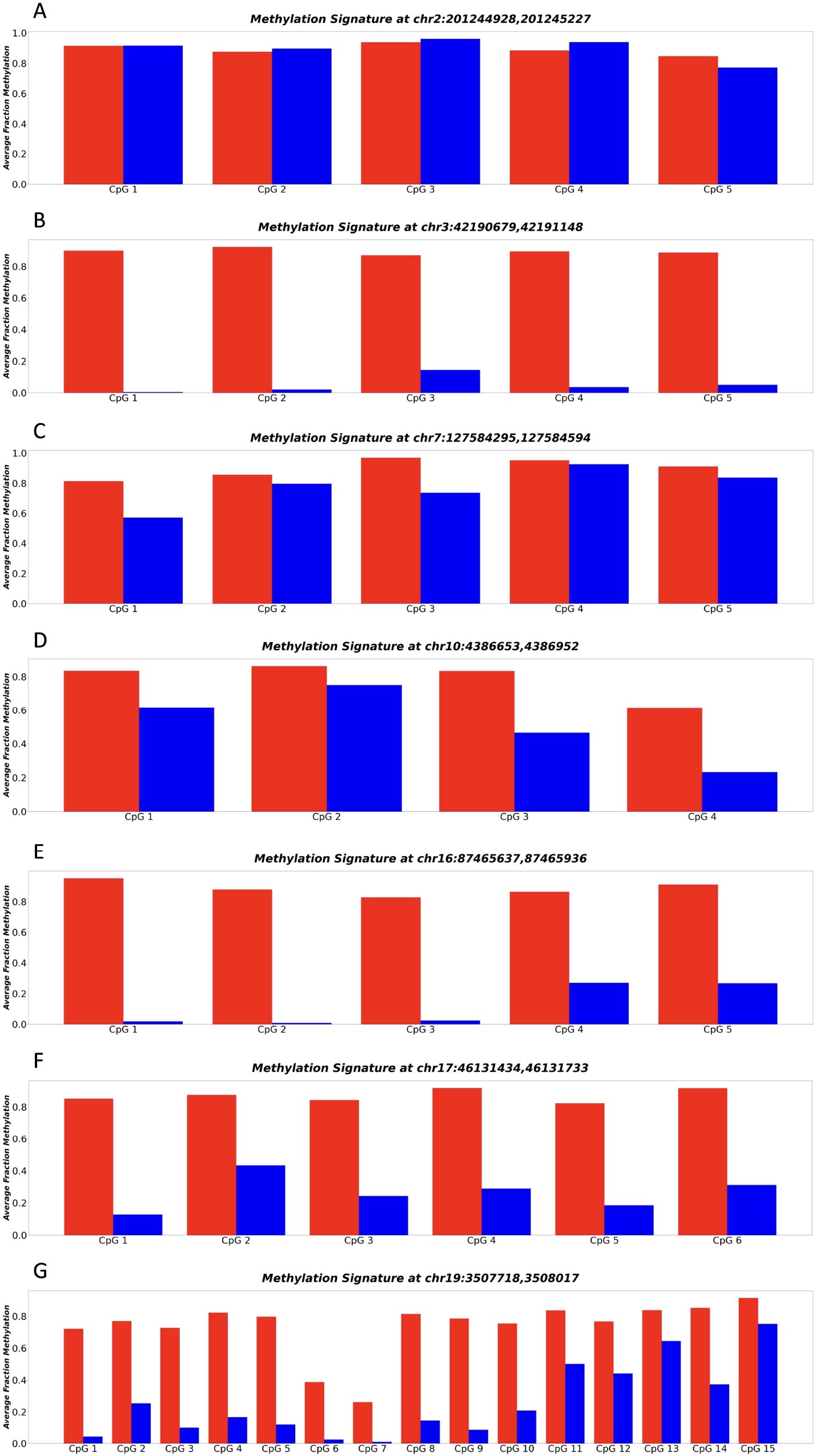
(A-G) represent the methylation patterns of seven sequenced loci in neuronal and blood plasma samples. Each panel displays the average fraction of methylation (y-axis) for individual CpGs (x-axis). Neuronal DNA shows complete unmethylation (blue regions), while blood plasma DNA exhibits near full methylation (red regions). These highly differential methylation patterns suggest the potential of these loci as robust biomarkers for neurodegenerative disease detection and monitoring.

### Identification of Neuron-Derived cfDNA in Patients

To identify neuron-derived cell-free DNA (cfDNA) in patients, targeted sequencing was performed at the locus chr3:42190679,42191148 on the bisulfite-converted DNA of our acquired dataset. Subsequently, we calculated the ratio of neuron-derived DNA for each sample. Strikingly, by using a conservative cutoff of 5% for the proportion of cfDNA derived from neurons in blood plasma, we achieved accurate identification of 100% of patients diagnosed with Alzheimer’s disease (AD) (Figure 4). In contrast, the young control cohort exhibited low or negligible levels of cfDNA derived from neurons, indicating the absence of neurodegeneration in this population.

**Figure 4:**
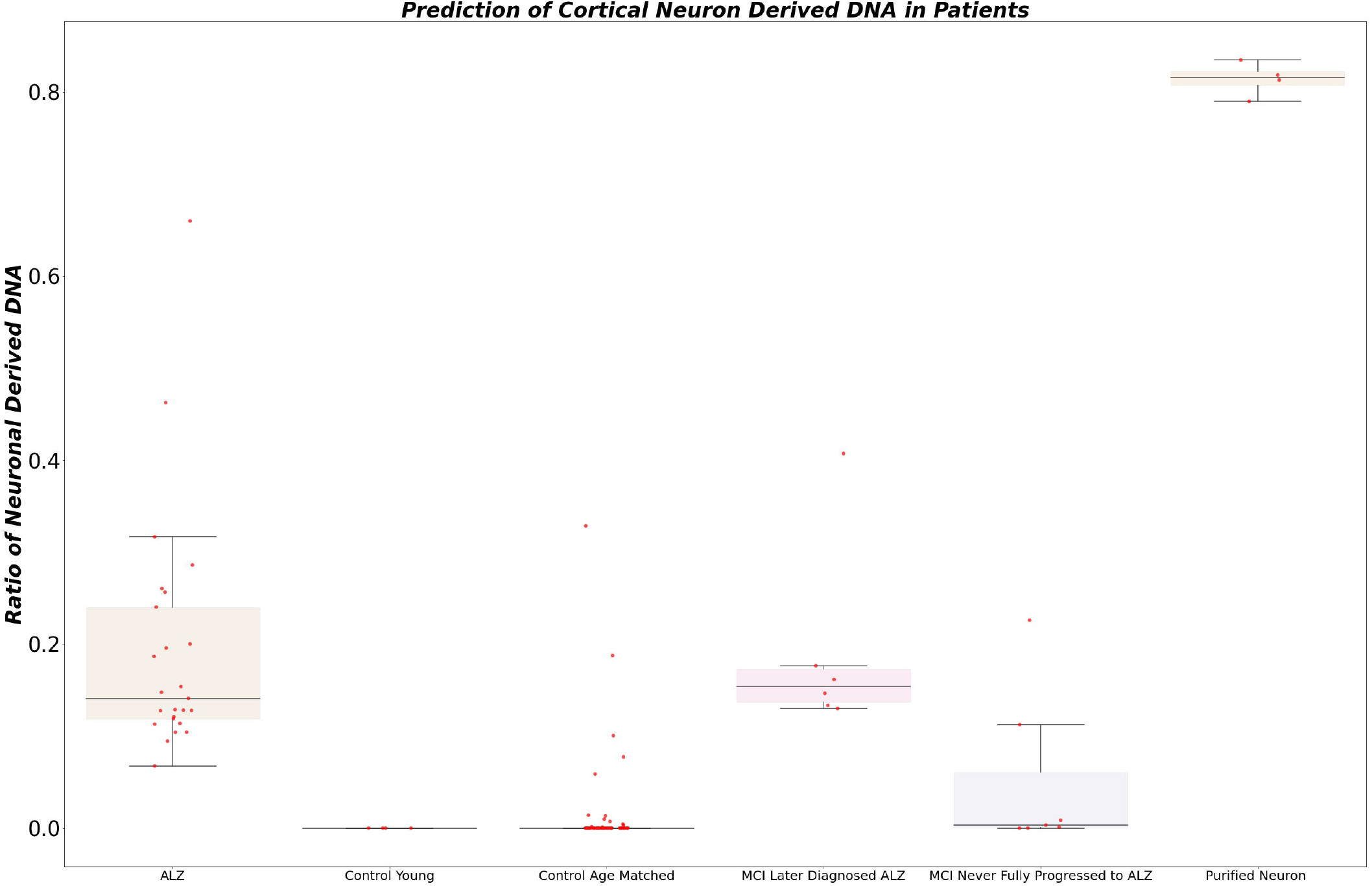
The figure illustrates the calculated ratio of neuron-derived DNA in blood plasma samples. Using a conservative cutoff of 5% for the proportion of cfDNA derived from neurons, the plot showcases the distribution of samples based on their neuron-derived cfDNA levels. The x-axis represents individual samples, while the y-axis denotes the percentage of cfDNA originating from neurons. The figure demonstrates the diagnostic potential of this ratio, accurately identifying patients diagnosed with AD and showing differentiation between healthy controls and individuals with mild cognitive impairment (MCI) who later progressed to AD.

Remarkably, all patients initially diagnosed with mild cognitive impairment (MCI) who later progressed to AD displayed elevated levels of neuron-derived cfDNA (>5% of total cfDNA). Among MCI patients who did not receive an Alzheimer’s diagnosis within 5 years, 75% exhibited normal or low levels of neuron-derived cfDNA, while 25% showed elevated levels.

In the aged healthy donor group, approximately 90% demonstrated normal levels of neuron-derived cfDNA, while approximately 10% exhibited elevated levels. Statistical analysis comparing the two groups of technical replicates revealed no significant difference, confirming the reproducibility and reliability of our experimental procedure (p-value 0.93). These compelling findings underscore the potential of our non-invasive blood-based approach for early detection and monitoring of neurodegenerative diseases, offering advantages of accessibility and ease of sample collection over traditional protein-based assays.

## Discussion

This study introduces a groundbreaking diagnostic approach for the early detection of neurodegenerative diseases by detecting neuron-derived cell-free DNA (cfDNA). Through a comprehensive analysis of Differential Methylation Regions (DMRs) between purified cortical neurons and blood plasma samples, we identified robust biomarkers that accurately distinguish neuron-derived cfDNA from non-neuron derived cfDNA, making them strong candidates for early disease detection. The use of cfDNA offers the advantage of convenient and minimally invasive sample collection compared to cerebrospinal fluid and tissue biopsies (11), enhancing the accessibility and ease of this diagnostic approach for patients. As a result, this method has the potential to predict the onset of various neurodegenerative diseases with high precision, revolutionizing the landscape of early diagnosis in the field of neuroscience.

In this study, we initially identified a significant number of DMRs (37,455) between purified neurons and blood plasma samples, indicative of a notable phenotypic difference. To optimize the practicality of this diagnostic approach, we further narrowed down the selection to 957 sites with a substantial mean methylation difference. From this subset, we selected the top 25 target sites and successfully validated their potential as biomarkers through primer design and amplification. Although this initial proof-of-concept study focused on a limited set of loci, future investigations should aim to develop a more comprehensive model that incorporates a larger number of loci to increase the robustness and accuracy of the diagnostic assay for Alzheimer’s disease and other forms of neurodegenerative disease.

Despite the promise of this diagnostic approach, certain limitations need to be addressed. One limitation is the reliance on a single specific locus to identify neuron-derived cfDNA. While we identified many potential loci that could serve as adequate predictors of neuron-derived cfDNA, our current approach only focused on a limited set of loci. This presents a problem as it increases the potential that this site is also correlated with other tissue types. To overcome this limitation, future research should explore the development of a more comprehensive assay that incorporates multiple loci, thus further enhancing the diagnostic accuracy.

Additionally, technical variance associated with PCR amplification and bisulfite conversion introduces the risk of batch effects (12). To ensure reliable and consistent results, appropriate controls and normalization procedures must be implemented to address these batch effects. In the long run, the development of an alternative assay that circumvents the need for bisulfite conversion and PCR would be beneficial, as it would likely eliminate batch effects and enhance the reproducibility of the diagnostic method. The use of nanopore sequencing, which enables the sequencing of native strand DNA while providing methylation data, holds promise as a potential platform to address these limitations (13).

Despite these challenges, our study has demonstrated the potential of utilizing cfDNA analysis as a valuable tool for pre-symptomatic detection and monitoring of neurodegenerative diseases. By employing a conservative cutoff of 5% for neuron-derived cfDNA, we achieved a remarkable ability to accurately identify patients with Alzheimer’s disease. Furthermore, the differentiation between MCI patients who later progressed to Alzheimer’s and those who did not showcases the potential of this diagnostic approach for early detection and prognostic assessment.

Notably, a small percentage of the aged control cohort exhibited elevated levels of neuron-derived cfDNA. This observation appears to align with the national average, indicating that 1 in 9 individuals above the age of 65 will have dementia (14). However, further research is needed to validate whether these individuals have indeed begun to show signs of cognitive decline.

In conclusion, this study highlights the advantages of utilizing cfDNA as a diagnostic tool for neurodegenerative diseases. By identifying differentially methylated regions between neurons and blood plasma, we have provided evidence for the feasibility of detecting neuron-derived cfDNA as an indicator of neurodegeneration. However, further research is required to expand the panel of predictive loci and develop improved methodologies to overcome current limitations. Nonetheless, this diagnostic approach holds promise for facilitating early detection, monitoring disease progression, and potentially enabling personalized therapeutic interventions for neurodegenerative disorders. With further development and validation, this innovative diagnostic strategy has the potential to significantly impact the field of neurodegenerative disease research and patient care.

## Funding

All funding was provided by Brigham Young University via a sponsored research agreement with the company Renew Biotechnologies.

## Author Contributions

Chad Pollard^1^ participated in the genesis of the research idea, sample processing, figure generation, and the writing of the manuscript. Kenneth Aston^2^ participated in manuscript drafting. Ben Emery^2^ participated in manuscript drafting. Jonathon Hill^1^ participated in the manuscript drafting, and technical optimization to obtain accurate sequencing results. Tim Jenkins^1*^ participated in the genesis of the research idea, obtaining the funding, sample acquisition, and final manuscript editing.

## Data Availability Statement

The microarray datasets analyzed for this study can be found in the Gene Expression Omnibus Repository under the accession numbers GSE108462 and GSE66351. All nanopore sequencing data is available upon request to the corresponding author.

## Notes

### Competing Interest Statement

All authors declare that they are affiliated with and have a financial relationship with either Renew Biotechnologies directly or its subsidiary company Wasatch Biolabs. Renew Biotechnologies funded this work via a sponsored research agreement with Brigham Young University and does retain the first right of refusal to any Intellectual Property that is created as a result of this research.

### Author Declarations

The PrecisionMed ethics committee gave ethical approval for this work. PrecisionMed also maintains that all samples acquired by their firm from outside institutions for redistribution have been acquired under the proper IRB approval. For access to patient consent reports please contact the corresponding author.

